# A Holistic Approach to Identification of Covid-19 Patients from Chest X-Ray Images utilizing Transfer Based Learning

**DOI:** 10.1101/2020.07.08.20148924

**Authors:** Taki Hasan Rafi

## Abstract

Novel coronavirus likewise called COVID-19 began in Wuhan, China in December 2019 and has now outspread over the world. Around 8 millions of individuals previously got influenced by novel coronavirus and it causes at any rate 500,000 deaths. There are just about 90,000 individuals contaminated by COVID-19 in Bangladesh too. As it is an exceptionally new pandemic infection, its diagnosis is challenging for the medical community. In regular cases, it is hard for lower incoming countries to test cases easily. RT-PCR test is the most generally utilized analysis framework for COVID-19 patient detection. However, by utilizing X-ray image based programmed recognition can diminish the expense and testing time. So according to handling this test, it is important to program and effective recognition to forestall transmission to others. In this paper, we attempt to distinguish COVID-19 patients by chest X-ray images. We execute different pre-trained deep neural system models, for example, Sequential, DenseNet121, ResNet152 and EfficientNetB4 to assess the most productive outcome. And aims to utilize transfer-based learning. We assess this outcome by AUC, where EfficientNetB4 has 0.997 AUC, ResNet50 has 0.967 AUC, DenseNet121 has 0.874 AUC and the Sequential model has 0.762 AUC individually. And EfficientNetB4 has achieved 98.86% accuracy.

## I. Introduction

A novel Coronavirus or COVID-19 is an infectious ailment brought about by a recently distinguished infection which is known to be just transmitted through a set of all animals yet as of late influenced people too. Since December 2019, various instances of “obscure viral pneumonia” identified with a nearby Seafood Wholesale Market was accounted for in Wuhan City, China [2]. A Novel coronavirus fit for tainting people was officially affirmed on January 6, 2020 [1]. As indicated by Nature, the spread of coronavirus ailment 2019 (COVID-19) is getting relentless and has just arrived at the important epidemiological measures for it to be announced a pandemic [19]. COVID-19 is an intense settled disease however it can likewise be lethal, with a 2% case casualty rate [6]. Like other coronaviral pneumonia, for example, a serious intense respiratory disorder brought about by coronavirus, COVID-19 can likewise prompt intense respiratory trouble condition [1].

There is a dire requirement for viable treatment. Current spotlight has been on the improvement of novel therapeutics, including antivirals and antibodies. Gathering proof recommends that a subgroup of patients with serious COVID-19 may have a cytokine storm condition [7]. The most widely recognized test method as of now utilized for COVID-19 determination is an ongoing converse interpretation polymerase chain response (RT-PCR) [8]. COVID-19 can cause intense heart injury. In the vast majority of the cases, the patients who have co-morbidity like diabetes, circulatory strain, coronary illness [10].

The side effects of these sicknesses resemble whatever another ordinary influenza which is a disadvantage of distinguishing the genuine influenced ones. The side effects can be demonstrated roughly in the middle of 14 days. As this COVID-19 is another infection for the clinical network, so still explicit treatment with respect to COVID-19 is difficult. There are some recognized side effects in regards to COVID-19, proposed by the World Health Organization (WHO). For example, high fever or mellow fever, hack, breathing problem, exhaustion, muscle or body throbs, migraine, loss of taste or smell, sore throat, clog or runny nose, spewing, diarrhoea. It straightforwardly influences the lung. X-Ray based images can assist us with knowing the lung condition so we can discover more COVID-19 cases as per the lung report. CT scan reports likewise can be utilized [3]. In spite of the fact that by far most of patients just have a typical, gentle type of sickness, around 15-20% of the patients fall into the serious gathering, which means they require helped oxygenation as a major aspect of treatment [20].

While it is about images-based problems, Deep Convolutional Neural Network can comprehend this all the more effectively these days. Deep neural-based frameworks can group images or related issues all the more precisely and productively by its condition of workmanship algorithmic strength. These are some enormous algorithms have been presented by deep learning researchers.

In this examination, We are to assess the viability of cutting edge pre-trained convolutional neural systems proposed by established researchers, with respect to their mastery in the programmed analysis of COVID-19 from thoracic X-ray images. We utilized pre-trained models, for example, DenseNet121, ResNet152, EfficientNetB4 and base convolutional model. Our assessment is dependent on AUC.

Further part of this paper-situated as related works, dataset, methodology, results, discussions and conclusion.

## II. Related Works

The novel coronavirus is another new disease in the field of the clinical network. Clinical researchers, just as deep learning specialists, are attempting to determine this issue. The fundamental test is to distinguish COVID-19 cases in a less measure of time and minimal cost. So the AI research network has come up to handle this test all the more proficiently. Less measure of works has been done as such far. In this segment, we go over some past and effective works with respect to this challenge from AI specialists.

A. Mangla et al. [9] have attempted to tackle COVID-19 case identification utilizing pre-prepared deep convolutional neural systems. Their model contains pre-prepared CheXNet, with a 121-layer Dense Convolutional Network (DenseNet) spine, trailed by a completely associated layer. They supplant CheXNet’s last classifier of 14 classes with our characterization layer of 4 classes, each with a sigmoid actuation to deliver the last yield. They wound up with a consequence of AUROC 0.9994 and precision of 87.2% in 4 class grouping. They named their model as CovidAID. El Asnaou et al. [11] have attempted to discover a few inquiries in regards to COVID-19 early recognition utilizing deep learning methods. They executed a few generally utilized deep learning structures, for example, VGG16, VGG19, MobileNet V2, Resnet50, DenseNet201, Inception ResNet V2 and Inception V3 in X-Ray just as Ct-Scan images. Where they infer that Inception ResNet V2 has performed superior to different architectures with a 92.18% accuray. I. Apostolopoulos et al. [12] have utilized pre-prepared deep learning models in their test. They tested in a dataset which contains 1427 X-Ray images. Where 700 images are typical pneumonia, 224 images with affirmed Covid-19 cases and 504 images of ordinary conditions. They utilized MobileNet v2, VGG19, Inception, Xception and Inception ResNet v2 designs. Where VGG 19 has given the best yield 98.75% accuracy in 2-class order. H. Abiyev et al. [13] conventional convolutional neural system to distinguish chest related ailment. They spoke to a correlation between the convolutional neural system, supervised back-propagation neural system and competitive neural system utilizing chest X-Ray images. Where the convolutional neural system has performed superior to different models. A. Abbas et al. [14] have actualized a tuned and altered deep neural system in X-ray images to distinguish COVID-19 cases all the more productively. They re-manufacture their model and named as DeTraC which contains 3 periods of layers. They built up this by utilizing ResNet-18 in backend and gets an accuracy of 95.12% in the X-Ray dataset. M. Rahimzadeh et al. [15] have actualized a connected of Xception and ResNet50V2 design to distinguish COVID-19 cases. In their trial, they utilized unbalanced X-Ray dataset. They observed numerous deep learning models look at the best result. The altered model which is a blend of Xception and ResNet50V2 has accomplished 91.40% accuracy on average. Naurin et al. [16] have executed convolutional neural systems, for example, Inception V3, Inception ResNetV3 and ResNet50 for the identification of COVID-19 cases by X-Ray images. They saw around 98% accuracy in pre-prepared ResNet50 model. Which is higher than Inception V3 model.

Considering all references, we attempt to handle this continuous COVID-19 detection issue by various deep learning procedures. We executed EfficientNetB4, ResNet50, DenseNet121 and base CNN model to legitimize which one performs better in this analysis.

## III. Dataset

In this investigation, we utilized and retrieved another arrangement of a dataset for the COVID-19 detection framework. It is accessible for theresearch community to battle against COVID-19 and quicken the exploration results. Later on, This dataset has been presented by Kaggle as an ongoing competition. The dataset contains a total of 5907 X-Ray images, where it has 5283 images for train purpose and 624 images for test purpose. It additionally has two classifications, for example, normal class and pneumonia class. Pneumonia class has four division, for example, SARS, COVID-19, ARDS and Streptococcus. The dataset can be downloaded from (https://github.com/ieee8023/covid-chestxray-dataset). The sample of the dataset has appeared in figure 1.

**Fig 1.**
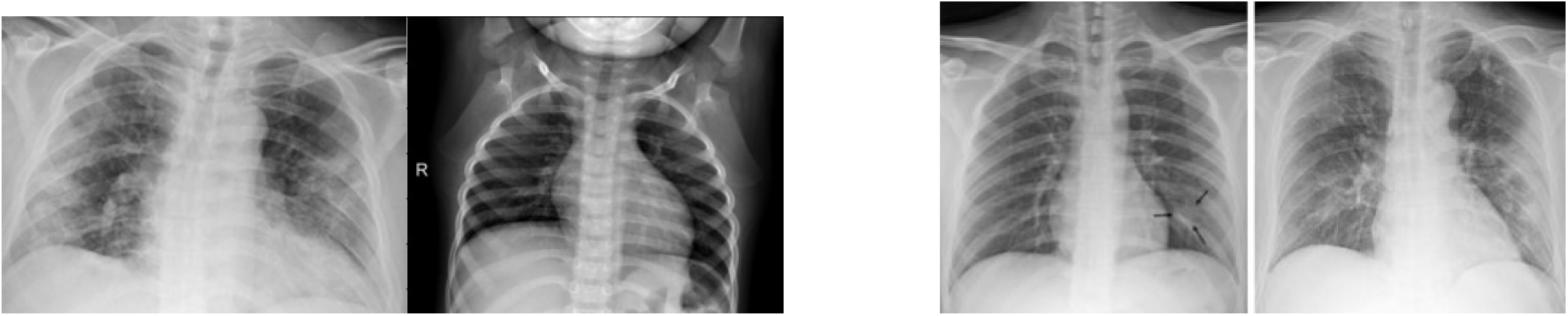
From left side, (a) Samples of normal case X-Ray and (b) Samples of COVID-19 case X-Ray.

**Fig 2.**
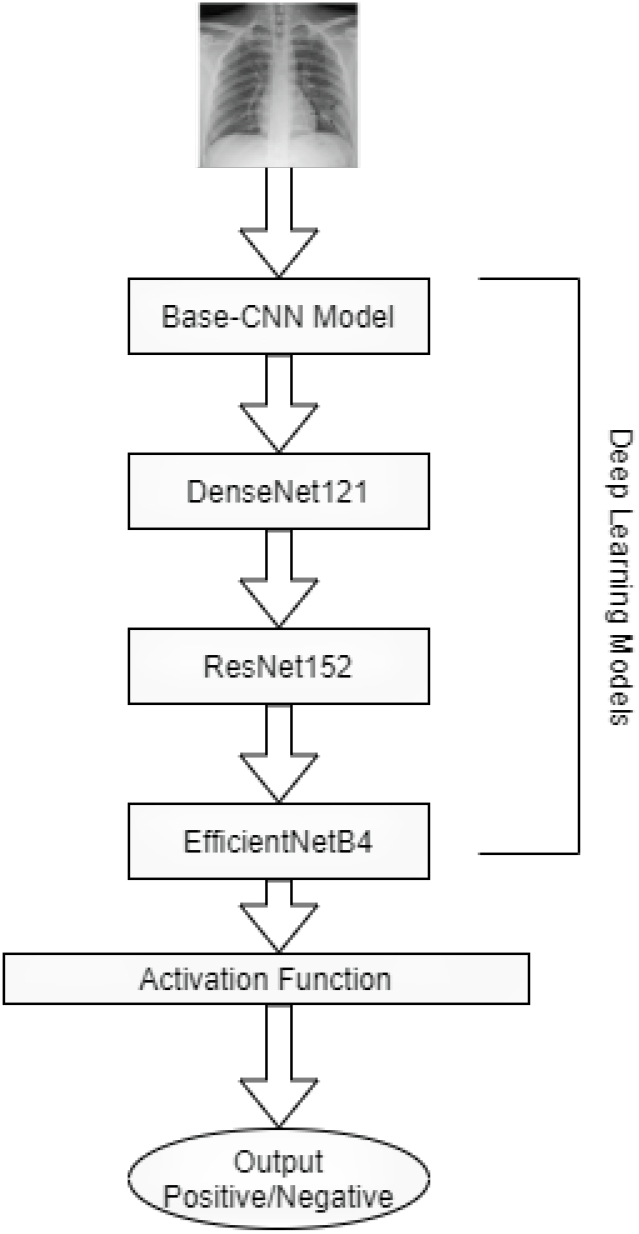
Block diagram of our methodology.

## IV. Methodology

In the dataset, we have 5907 images to investigate the examination. Where the dataset has a few arrangements of images, for example, Normal, SARS, COVID-19, ARDS and Streptococcus. Be that as it may, in this analysis our primary goal to identify COVID cases. To identify all the more proficiently, in this manner we are going to build a model which separates between normal case, Pneumonia and COVID-19 Cases. We additionally lessen the classification number to two. Later on, we applied different pre-trained deep learning models, for instance, Base Convolutional Neural Network, DenseNet121, ResNEt50 and EfficientNetB4 to distinguish COVID-19 cases and to locate the best exact outcome as indicated by the individual exhibitions.

### A. Visualization

We attempted to visualize and separate our characterization images for better understanding. Therefore, we envisioned our data by utilizing image histogram. An images histogram is a sort of histogram that goes about as a graphical portrayal of the apparent circulation in an advanced images. It plots the number of pixels for each apparent worth. By taking a gander at the histogram for a particular image a watcher will have the option to pass judgment on the whole apparent appropriation initially. Here figure 3 expresses the visualized images.

**Fig 3.**
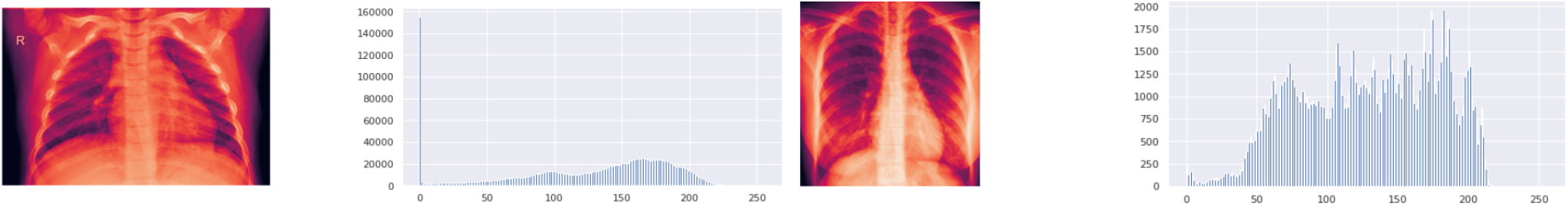
From left side, (a) Histogram of normal case and (b) Histogram of COVID-19 case.

**Fig 4.**
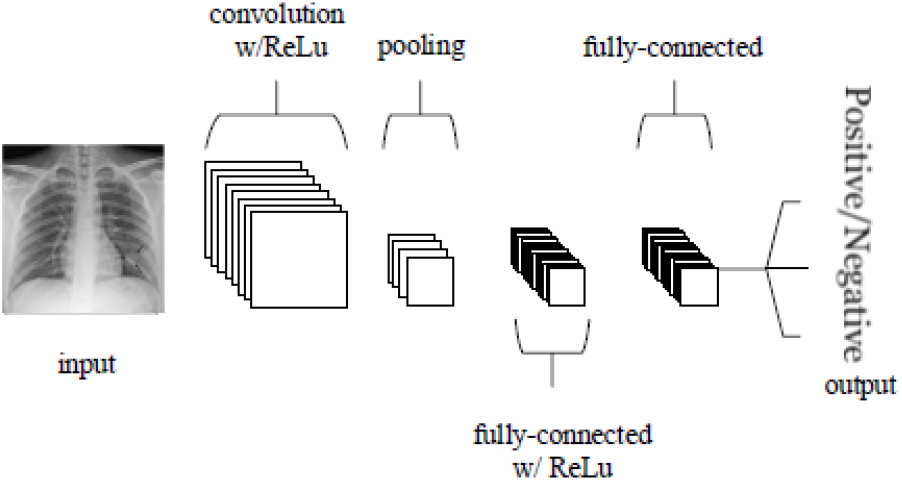
CNN Architecture.

### B. Base Convolutional Neural Network

Convolutional Neural Networks are practically equivalent to conventional Artificial Neural Networks in that they are included neurons that self-streamline through learning [18]. It fundamentally center around the premise that the info will be involved images. This centers the engineering to be set up in a manner to best suit the requirement for managing the particular sort of data. There are a few functionalities to explain Convolutional Neural Network more briefly. As found in various kinds of Artificial Neural Network, the information layer will hold the pixel estimations of the image. Convolutional layers will choose the yield of neurons of which are related to close-by regions of the commitment through the check of the scalar the thing between their heaps and the area related to the data volume.

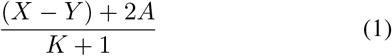

Here, X is the representation of input volume size (height x weight x depth), Y stands for the receptive field size, A stands for the size of zero padding and K stands for stride. The reviewed straight unit hopes to apply an elementwise inception capacity or initiation work, for instance, sigmoid to the yield of the sanctioning made by the past layer. Pooling layers will by then simply perform down-testing along the spatial the dimensionality of the given information, further reducing the amount of limits inside that incitation. completely associated layers will by then play out comparative commitments found in standard Artificial Neural Networks and try to convey class scores from the institutions, to be used for portrayal. It is moreover suggested that ReLu may be used between these layers, as to improve execution. Zero-padding is the basic procedure of cushioning the outskirt of the information and is a compelling strategy to give further control with regards to the dimensionality of the yield volumes. Boundary sharing chips away at the supposition that in the event that one area highlight is helpful to register at a set spatial area, at that point, it is probably going to be valuable in another locale. In the event that we compel every individual initiation map inside the yield volume to similar loads and predisposition, at that point we will see a huge decrease in the quantity of boundaries being created by the convolutional layer [18].

### C. Densely Connected Convolutional Network

Densely Connected Convolutional Network (DenseNet) interfaces each layer to each other layer in a feed-forward design [17]. While convolutional neural systems with N layers have N associations—one between each layer and its ensuing layer. DenseNets have a few convincing points of interest: they lighten the vanishing gradient issue, fortify element spread, empower include reuse, and significantly diminish the quantity of boundaries. It has better accuracy than ResNet in object recognition [17]. DenseNets are worked from thick squares and pooling activities, where each thick square is an iterative connection of past element maps. This design can be viewed as an augmentation of ResNets [5], which performs an iterative summation of past component maps. In any case, this little change makes them intrigue suggestions such as, boundary proficiency, DenseNets are more efficient in the boundary use.

Understood profound oversight, DenseNets perform profound management on account of short ways to all component maps in the design and highlight reuse, all layers can without much of a stretch access their first layers making it simple to reuse the data from recently figured element maps. The attributes of DenseNets make them an awesome fit for the semantic division as they normally actuate skip associations and multi-scale management. Fully connected DenseNets are worked from a downsampling way, an upsampling way and skip associations. Skip associations help the upsampling way recoup spatially point by point data from the downsampling way, by reusing highlights maps. The objective of our model is to additionally misuse the component reuse by broadening the more refined DenseNet engineering while at the same time maintaining a strategic distance from the element blast at the upsampling way of the system [17]. In this investigation, we utilized pretrained DenseNet121 design to actualize in our dataset.

### D. Residual Network

Residual Network has been created and acquainted by Microsoft Research with handle image recognition all the more without any hurdle. ResNet has about 3.57% less error than VGG Net [5]. It has around 152 layers top to bottom, which is eight multiple times higher than VGGNet design. Its architecture has been inspired by VGG Nets architecture. We are meaning the mapping as *H*(*a*), another non-direct mapping can be communicated as *F* (*a*) = *H*− (*a*) *a*, the primary mapping can be communicated as *F* (*a*) + *a*. We receive lingering figuring out how to each couple of stacked layers. The structure square can be defined as [5]:

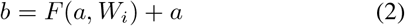

Here, a and b can be considered as the vectors. The function *F* (*a, Wi*) is considered as figuring out how to outline. The elements of a and F must be the equivalent. On the off chance that we can not figure out how to do as such, we can include a projection vector into the detailing. The projection must be straight and, for example, *W*_*s*_. Yet, we will appear by tests that the character mapping is adequate for tending to the debasement issue and is efficient, and accordingly, *W*_*s*_ is possibly utilized when coordinating measurements. It has two plan rule and scarcely followed by VGG Net. The principles are: It has multiple times 3 convolutional layers, for a similar yield highlight map size, the layers have a similar number of channels and if the component map size is split, the quantity of channels is multiplied in order to save the time multifaceted nature per layer [5]. The downsampling stride must be 2. In this analysis, we utilized pre-trained ResNet152 architecture.

### E. EficientNet

EfficientNet has been created and presented by Mingxing Tan, Staff Software Engineer at Google. EfficientNet is a systematical model scaling and distinguishes that cautiously adjusting system profundity, width, and goals can prompt better performance [4]. It is propelled by ResNet and MobileNet, and scalling up or down to legitimize better exactness.This is a compound demonstrating framework. There are numerous approaches to scale a ConvNet for various asset limitations: ResNet [5] can be downsized for example ResNet-18 or up e.g., ResNet-200 by altering system profundity or the quantities of layers.A convolutional neural layer can be detailed as *B*_*i*_ = *F*_*i*_(*A*_*i*_), where, *B*_*i*_ is the yield tensor, *A*_*i*_ is the information tensor and *F*_*i*_ is the employable capacity. tensor shape (*X*_*i*_, *Y*_*i*_, *Z*_*i*_) where *X*_*i*_ and *Y*_*i*_ are spatial measurements and *Z*_*i*_ is the channel measurement. There are three significant boundaries to consider for scaling reason, for example, profundity, width and goals. Scaling system profundity is the most well-known way utilized by numerous convolutional systems. The instinct is that more profound convolutional systems can catch more extravagant and progressively complex highlights, and sum up well on new errands. Notwithstanding, more profound systems are additionally progressively hard to prepare because of the disappearing inclination issue. The genuine errand of this model is to scaling the profundity, width and goals all the more productively to change the assignment prerequisite. goals with a lot of fixed scaling coefficients. For instance, on the off chance that we need to utilize 2^*n*^ times progressively computational assets, at that point we can just build the system profundity by *α*^*n*^, width by *β*^*n*^, and image size by *γ*^*n*^, where *α, β, γ* are consistent coefficients controlled by a little lattice search on the first little model. In this analysis, we utilized pre-trained EfficientNetB4 model for COVID-19 for detecting purpose.

### F. Activation Function

Actuation functions are numerical conditions that decide the yield of a neural system. The capacity is appended to every neuron in the system and decides if it ought to be actuated or not, founded on whether every neuron’s input is significant for the model’s expected output. In this investigation, we utilized two individual activation functions, for example, Sigmoid [26] and ReLU [23] activation function. We utilized Sigmoid in Base CNN model and ReLU in rest of different models as initiation work.

- **Sigmoid:** The sigmoid activation function is here and there alluded to as the strategic capacity or crushing capacity in some literatures [26]. The Sigmoid is a non-direct enactment work utilized for the most part in feedforward neural systems.

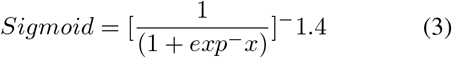 The sigmoid capacity shows up in the yield layers of the deep learning structures, and they are utilized for anticipating likelihood-based yield and has been applied effectively in binary characterization issues, demonstrating strategic relapse undertakings just as other neural system areas [26].
- **ReLU:** Rectified Linear Units (ReLU) as the arrangement work in a deep neural system [23]. Customarily, ReLU is utilized as an actuation work in deep neural systems, with Softmax work as their arrangement work. It works by thresholding values at 0, model *f* (*a*) = *max*(0, *a*). Basically, it yields 0 when *a <* 0, and then again, it yields a straight capacity when *a*0. ReLU is not just as an activation function in each concealed layer of a neural system yet additionally as the grouping capacity at the last layer of a system. Thus, the anticipated class for ReLU classifier would be 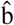 [23],

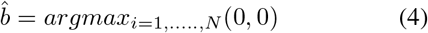

### G. Optimizer

The optimizer is a robust algorithm that helps to reduce the loss of a deep neural system by changing some attributes such as learning rate and changing weight and enhance the overall performance of the system. An optimizer can improve the performance of a neural system. It is essential to use an optimizer to reduce loss functions. In this experiment, we used two extensively used optimizers such as Stochastic Gradient Descent and Adam. We used Adam in EfficientNetB4 algorithm and ResNet152, on the other hand, we used Stochastic Gradient Descent (SGD) in DenseNet121 and Base CNN model.

- **Stochastic Gradient Descent:** Stochastic Gradient Descent is widely utilized optimizer, much of the time, it is utilized in traditional CNN model to streamline [25]. It is an updated form of Batch SGD. SGD gets rid of this repetition by performing each update in turn. It is subsequently generally a lot quicker and can likewise be utilized to learn on the web. SGD performs visit refreshes with a high change that prompt the target capacity to change intensely.
- **Adam:** Adam is an extensively utilized optimizer, which is 1st order gradient-based optimizer. It is a strategy for proficient stochastic advancement that just requires first-request slopes with little memory prerequisite. The technique registers individual versatile taking in rates for various boundaries from evaluations of first and second snapshots of the angles, the name Adam is gotten from versatile second estimation [24]. This can be formulated as:

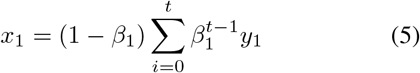

## V. Performance Matrix

We evaluated our models by AUC, accuracy, precision, specificity and sensitivity.

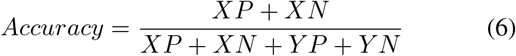

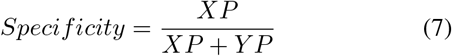

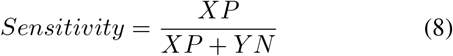

Here, XP and XN denote as true positive and true negative, YP and YN denote as false positive and false negative respectively.

## VI. Results and Discussions

In this analysis, we utilized a few convolutional neural system models to empower better outcome. We executed EfficientNEtB4, ResNet152, DenseNet121 and Base convolutional neural Network model to identify COVID-19 cases all the more proficiently. There are a few works have been done before in an exceptionally brief timeframe to handle this obstacle by colossal scientists. Different specialists have created gathering algorithms for identifying COVID-19 cases. In our examination, EfficientNetB4 has performed better.

It has 98.86% accuracy and 0.996 AUC. Different models, for example, ResNet152, DenseNet121 and Base CNN have additionally performed well. We set the epochs to 20 in every examination. Be that as it may, Base CNN has the most reduced accuracy of 84.50% where ResNet152 has 97.31% and DenseNet121 has an accuracy of 96.50%. We executed the sigmoid activation function in Base CNN model and ReLU activation function in the remainder of the models. Then again, we utilized stochastic gradient descent (SGD) optimizer in Base CNN model and DenseNet121 model. We likewise utilized Adam optimizer in EfficientNetB4 and resNet152 model. In table 1. we have demonstrated the presentation examination of each algorithm. In figure 5, we have shown performances of every algorithm with AUC, training loss, validation loss and validation AUC. And in figure 6, we have shown EfficientNetB4’s model accuracy by increasing epochs and model loss in order to increase epochs.

**TABLE 1.** Performance Analysis of Every Algorithm (%)

| Algorithm | AUC | Accuracy | Specificity | Sensitivity |
| --- | --- | --- | --- | --- |
| Base CNN Model | 0.762 | 84.50 | 81.43 | 88.29 |
| DenseNet121 | 0.874 | 96.50 | 92.66 | 93.28 |
| ResNet152 | 0.967 | 97.31 | 97.78 | 96.12 |
| **EfficientNetB4** | **0.997** | **98.87** | **99.46** | **98.77** |

**Fig 5.**
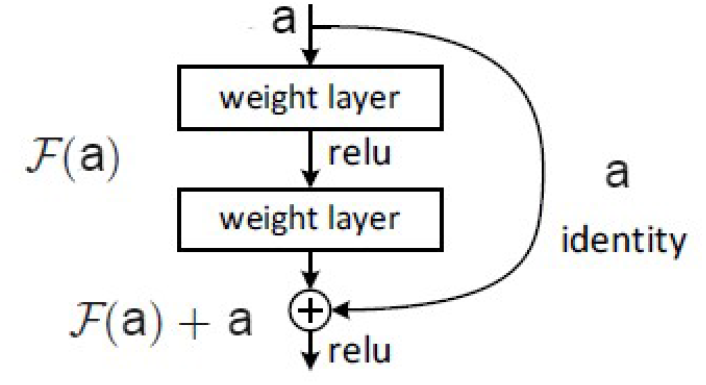
ResNet block diagram [5].

**Fig 6.**
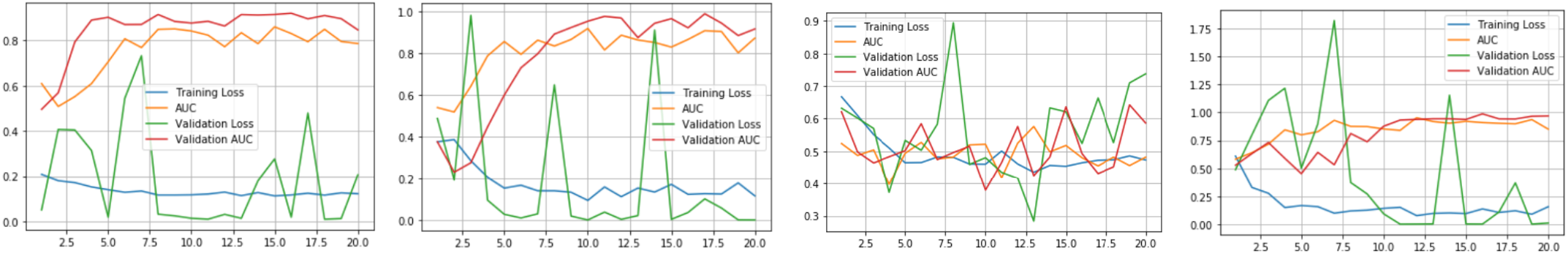
From left side, (a) Base CNN performance graph; (b) ResNet152 performance graph; (c) DenseNet121 performance graph and (d) EfficientNetB4 performance graph.

**Fig 7.**
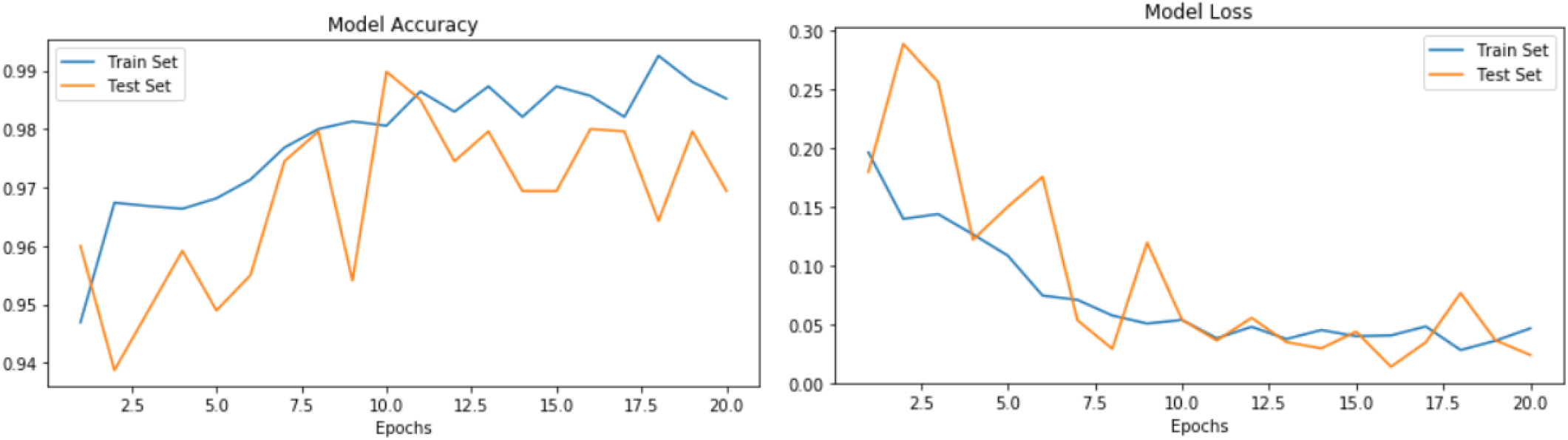
From left side, (a) Model accuracy of EfficientNetB4 model and (b) Model loss of EfficientNetB4 model.

Finally, because of excellent outcome in chest X-Ray images, we propose and concoct the choice that EfficientB4 model can be utilized for additional identifying limit with regards to clinical network to handle this hardest time for the world.

## VII. Conclusion

Conclusion of COVID-19 is basic to follow the influenced individuals and limit the transmission as it is a viral disease. RT-PCR method is costly and needs more time to detect COVID-19 cases. These days medical images preparing is one of the fundamental assignments for the scientists to foresee or recognize infections all the more productively. So as referenced, we attempted to handle COVID-19 discovery issue by utilizing medical images, all the more explicitly chest X-Ray images. In this investigation, We have actualized a few pre-tranied deep convolutional neural networks, for example, Base CNN, DenseNet121, ResNet152 and EfficientNetB4 in chest X-Ray dataset. Convolutional neural systems can think of an effective and powerful result than any conceivable way. In our investigation, EfficientNetB4 has performed superior to different models with an accuracy of 98.86% just as containing higher AUC of 0.996. Then again, ResNet152 has likewise performed well with accuracy and AUC of 97.31% and 0.967 separately. Different models, for example, DenseNet121 and Base CNN have an accuracy of 96.50% and 84.50%. So in rundown, we would recommend and proposed EfficientNetB4 for additional headway of distinguishing COVID-19 cases by utilizing X-Ray images. For future improvement, we need a more sophisticated dataset with more amount of images to train our model for better outcomes.

## Data Availability

https://github.com/ieee8023/covid-chestxray-dataset

## References

[1] F. Pan et al., “Time Course of Lung Changes at Chest CT during Recovery from Coronavirus Disease 2019 (COVID-19),” Radiology, vol. 295, no. 3, pp. 715–721, 2020, doi:10.1148/radiol.2020200370.

[2] T. Ai, Z. Yang, and L. Xia, “Correlation of Chest CT and RT-PCR Testing in Coronavirus Disease,” Radiology, vol. 2019, pp. 1–8, 2020, doi:10.14358/PERS.80.2.000.

[3] Charmain Butt, “covid19: Deep Learning System to Screen Coronavirus Disease 2019 Pneumonia,” 2020, [Online]. Available: http://arxiv.org/abs/2002.09334.

[4] M. Tan and Q. V. Le, “EfficientNet: Rethinking model scaling for convolutional neural networks,” 36th Int. Conf. Mach. Learn. ICML 2019, vol. 2019-June, pp. 10691–10700, 2019.

[5] K. He, X. Zhang, S. Ren and J. Sun, “Deep Residual Learning for Image Recognition,” in Prac. 2016 IEEE Conference on Computer Vision and Pattern Recognition (CVPR), Las Vegas, NV, 2016, pp. 770–778, doi:10.1109/CVPR.2016.90.

[6] Z. Xu et al., “Pathological findings of COVID-19 associated with acute respiratory distress syndrome,” Lancet Respir. Med., vol. 8, no. 4, pp. 420–422, 2020, doi:10.1016/S2213-2600(20)30076-X.

[7] P. Mehta, D. F. McAuley, M. Brown, E. Sanchez, R. S. Tattersall, and J. J. Manson, “COVID-19: consider cytokine storm syndromes and immunosuppression,” Lancet, vol. 395, no. 10229, pp. 1033–1034, 2020, doi:10.1016/S0140-6736(20)30628-0.

[8] T. Ozturk, M. Talo, E. Azra, U. Baran, and O. Yildirim, “Automated detection of COVID-19 cases using deep neural networks with X-ray images,” no. January, 2020.

[9] A. Mangal et al., “CovidAID: COVID-19 Detection Using Chest X-Ray,” no. May, 2020, [Online]. Available: http://arxiv.org/abs/2004.09803.

[10] Y. Y. Zheng, Y. T. Ma, J. Y. Zhang, and X. Xie, “COVID-19 and the cardiovascular system,” Nat. Rev. Cardiol., vol. 17, no. 5, pp. 259–260, 2020, doi:10.1038/s41569-020-0360-5.

[11] K. El Asnaoui and Y. Chawki, “Using X-ray images and deep learning for automated detection of coronavirus disease,” J. Biomol. Struct. Dyn., vol. 0, no. 0, pp. 1–12, 2020, doi:10.1080/07391102.2020.1767212.

[12] I. D. Apostolopoulos and T. A. Mpesiana, “Covid-19: automatic detection from X-ray images utilizing transfer learning with convolutional neural networks,” Phys. Eng. Sci. Med., no. 0123456789, pp. 1–6, 2020, doi:10.1007/s13246-020-00865-4.

[13] R. H. Abiyev and M. K. S. Ma’aitah, “Deep Convolutional Neural Networks for Chest Diseases Detection,” J. Healthc. Eng., vol. 2018, 2018, doi:10.1155/2018/4168538.

[14] A. Abbas, M. M. Abdelsamea, and M. M. Gaber, “Classification of COVID-19 in chest X-ray images using DeTraC deep convolutional neural network,” 2020, [Online]. Available: http://arxiv.org/abs/2003.13815.

[15] M. Rahimzadeh and A. Attar, “A modified deep convolutional neural network for detecting COVID-19 and pneumonia from chest X-ray images based on the concatenation of Xception and ResNet50V2,” Informatics Med. Unlocked, vol. 19, p. 100360, 2020, doi:10.1016/j.imu.2020.100360.

[16] Narin, Ali Kaya, Ceren Pamuk, Ziynet. (2020). Automatic Detection of Coronavirus Disease (COVID-19) Using X-ray Images and Deep Convolutional Neural Networks.

[17] G. Huang, Z. Liu, L. Van Der Maaten, and K. Q. Weinberger, “Densely connected convolutional networks,” Proc. - 30th IEEE Conf. Comput. Vis. Pattern Recognition, CVPR 2017, vol. 2017-January, pp. 2261–2269, 2017, doi:10.1109/CVPR.2017.243.

[18] K. O’Shea and R. Nash, “An Introduction to Convolutional Neural Networks,” no. November, 2015, [Online]. Available: http://arxiv.org/abs/1511.08458.

[19] A. Remuzzi and G. Remuzzi, “COVID-19 and Italy: what next?,” Lancet, vol. 395, no. 10231, pp. 1225–1228, 2020, doi:10.1016/S0140-6736(20)30627-9.

[20] S. Tian, W. Hu, L. Niu, H. Liu, H. Xu, and S. Y. Xiao, “Pulmonary Pathology of Early-Phase 2019 Novel Coronavirus (COVID-19) Pneumonia in Two Patients With Lung Cancer,” J. Thorac. Oncol., vol. 15, no. 5, pp. 700–704, 2020, doi:10.1016/j.jtho.2020.02.010.

[21] A. I. Khan, J. L. Shah, and M. M. Bhat, “CoroNet: A Deep Neural Network for Detection and Diagnosis of COVID-19 from Chest X-ray Images,” Comput. Methods Programs Biomed., p. 105581, 2020, doi:10.1016/j.cmpb.2020.105581.

[22] C. Tan, F. Sun, T. Kong, W. Zhang, C. Yang, and C. Liu, “A survey on deep transfer learning,” Lect. Notes Comput. Sci. (including Subser. Lect. Notes Artif. Intell. Lect. Notes Bioinformatics), vol. 11141 LNCS, pp. 270–279, 2018, doi:10.1007/978-3-030-01424-727.

[23] Agarap, Abien Fred. (2018). Deep Learning using Rectified Linear Units (ReLU).

[24] Kingma, Diederik Ba Jimmy. (2014). Adam: A Method for Stochastic Optimization. in Prac. International Conference on Learning Representations, Banff, Canada.

[25] S. Ruder, “An overview of gradient descent optimization algorithms.” 2016.

[26] Nwankpa, Chigozie Ijomah, Winifred Gachagan, Anthony Marshall, Stephen. (2018). Activation Functions: Comparison of trends in Practice and Research for Deep Learning.

